# Multiplexed micronutrient, inflammation, and malarial antigenemia assessment using a plasma fractionation device

**DOI:** 10.1101/2021.11.04.21265891

**Authors:** Eleanor Brindle, Lorraine Lillis, Rebecca Barney, Pooja Bansil, Francisco Arredondo, Neal E. Craft, Eileen Murphy, David S. Boyle

## Abstract

Collecting, processing, and storing blood samples for future analysis of biomarkers can be challenging when performed in resource limited environments. The preparation of dried blood spots (DBS) from heel or finger stick collection of whole blood is a widely used and established method. DBS pose less risk of infection from blood borne pathogens, do not require immediate specimen processing and tolerate a wider range of storage temperatures, and are easier to ship. As such, DBS are commonly used in large-scale surveys to assess infectious disease status and/or micronutrient status in vulnerable populations. Recently, we reported that DBS can be used with a multiplexed immunoassay, the Q-plex™ Human Micronutrient 7-plex Array (MN 7-plex). This tool can simultaneously quantify seven protein biomarkers related to micronutrient deficiencies (iodine, iron and vitamin A), inflammation and malarial antigenemia using plasma or serum. Serum ferritin, a key iron biomarker, cannot be measured from DBS due to red blood cell (RBC) ferritin confounding the results. In this study, we demonstrate the performance of a simple and rapid blood fractionation tool that passively separates serum from cellular components via diffusion through a membrane into a plasma collection disc (PCD) to produce plasma spots. We evaluated the concordance of MN 7-plex analyte concentrations from matched panels of eighty-eight samples of PCD, DBS, and wet plasma prepared from anticoagulated venous whole blood. The results show high correlation between eluates from PCD and DBS and wet plasma for each analyte. Serum ferritin measures from the PCD eluates were highly correlated to wet plasma samples. This suggests that surveillance for iron deficiency may be improved over the current methods restricted to only measuring sTfR in DBS as when used in combination with the MN 7-plex, all seven biomarkers can be simultaneously measured using PCDs.

## Introduction

Micronutrient deficiency (MND) has been described as hidden hunger and an estimated 2 billion people are affected globally [1] with Iodine, Iron, and vitamin A representing three of the four primary MNDs of global significance [2-4]. Sufficient concentrations of key micronutrients (MN) are required for normal development of the fetus *in utero*, infants, and young children. MND can impair cognitive, immune, ocular function and physical health and development. Fetuses, young children, and women of reproductive age in are particularly at risk from MND and the global burden is greatest in low- and middle-income countries (LMICs) that are the least equipped to routinely assess MN status [5].

There are low cost and effective solutions to mitigate the effects of MND including dietary initiatives, fortification of common foodstuffs (e.g. salt or flour) or directly via supplementation [6-8]. The challenge to nutrition programs and researchers in LMICs is to effectively perform population surveillance to identify groups at most risk of MND before its effects manifest. Baseline data can identify the scale of MND and groups at greatest risk so that effective interventions can be introduced. Interventions can then be monitored via ongoing surveillance at scheduled intervals to establish their impact.

There is general consensus on a panel of blood-based biomarkers that are both informative on iodine, iron or vitamin A status [9-13], and practical for use in population-level surveillance. These act as surrogates to more direct MN measurement methods which involve challenging specimen collection (e.g. bone marrow aspirates for iron measurement [14]) or require highly skilled users and complex equipment (e.g. high pressure liquid chromatography for serum retinol analysis [15]). The blood specimens for these tests can be collected by venipuncture or less invasive methods, such as finger- or heel-stick capillary blood collection. Venous blood collection offers the advantage of much larger sample volumes and reduced chance of bias resulting from the capillary blood collection technique (e.g. milking fingers), but venous samples are more difficult to collect, particularly from children, and more challenging to transport, process, and store [16]. Serum or plasma from whole blood must be rapidly processed to avoid hemolysis and must be kept under cold chain to prevent decay of the biomarkers. A perception of greater pain associated with venipuncture versus finger stick collection may also negatively influence participation in studies [17]. Blood and its derivatives pose biohazard risks, and handling should only be performed by staff with certified training for blood borne pathogens [18]. Therefore, the scaled collection of venous blood adds significant logistical, financial and technical challenges particularity in low resource settings where the need for effective MND surveillance is greatest.

The use of DBS as an alternative sample type meets many of the challenges presented with venous whole blood. Blood collection of up to 250 µL from a heel or finger prick via sterile lancet can be performed by less skilled users [28]. The blood is spotted on paper card and left to dry before being stored in a sealed pouch with a desiccant to eliminate moisture. The preparation of a sample takes less time, needs no ancillary equipment (e.g. centrifuge or refrigeration) and uses less materials and consumables [16,19,20]. Once dried, the sample has greatly reduced biohazard risk during sample handling and processing. DBS cards have the further advantages of being relatively small requiring significantly less space in freezers during long-term storage, and as many biomarkers are stable in DBS at a broader range of temperatures, they can be shipped using gel packs instead of dry ice, or even out of cold chain [19,28-31].

A primary challenge in using DBS is that the sample quantity is limited, with a typical collection yielding five spots of whole blood with approximately 50 μL each. Eluting DBS from filter paper results in dilution of the specimen, so assays must be capable of quantifying low concentrations. We recently highlighted that low concentration biomarkers such as thyroglobulin (Tg, µg/L) could be quantified from DBS eluates using the Q-Plex™ Human Micronutrient Array (Quansys Biosciences, Logan, Utah, USA) [21,22]. The Q-plex is a multiplexed immunoarray that enables the simultaneous quantification of biomarkers to deficiencies in iodine, iron, and vitamin A, in addition to inflammatory biomarkers and *Plasmodium falciparum*, from a single small volume of liquid serum or plasma [21].

While this evaluation found acceptable agreement between DBS and plasma for most analytes, using DBS for iron deficiency assessment is challenging, because ferritin levels are grossly elevated by the co-elution of serum and red blood cell ferritin from lysed RBCs. Therefore, sTfR is the only iron biomarker that can be measured from DBS [22,23]. While sTfR is one of the recommended biomarkers for iron status assessment, we and other groups have reported challenges in harmonizing absolute sTfR values derived from different immunoassay methods [19,21,24-26]. The ratio of sTfR to serum ferritin has been found to be a better indicator of iron deficiency than either analyte alone [27]. As such, we hypothesize that a parallel measurement of ferritin, inflammatory biomarkers and sTfR may give greater confidence in establishing iron deficiency status. A variety of simple devices have been described or developed to passively fractionate plasma from whole blood, primarily to enable more accurate determination of viral load testing of people living with HIV [28,29].

In this manuscript we describe an evaluation of use of the Q-plex™ micronutrient 7-Plex assay (hereafter MN 7-plex) with matched sets of plasma, eluates of DBS, and eluates from a prototype of the plasma collection disc (PCD), a passive blood fractionation device (a commercial version, the DriVive is now offered by ViveBio LLC, Alpharetta, GA, USA) [30]. In particular, we evaluate whether the PCD allows reliable measurement of ferritin using a method with the same logistical advantages as DBS. We also evaluate performance of the MN 7-plex using DBS and plasma prepared from heparinized whole blood samples as opposed to EDTA blood used in an earlier DBS study [22].

## Materials and methods

### Collection of whole blood specimens

Specimens of ∼8 mLs of human whole blood were procured from a commercial vendor, BioIVT (Westbury, New York). The WIRB protocol number for the collection of 80 blood samples was 20161665 and all specimens were obtained only after signed consent by each participant. The PATH Office of Research Affairs deemed the use of these samples to be non-human subjects research.

### Preparation of DBS, PCD and plasma panels

A panel of 80 heparinized anticoagulated whole blood samples (40 adult male, 40 adult female) was procured. The whole blood was delivered to the PATH laboratory the next day, and processed upon receipt to prepare three sample types (DBS, PCD, and plasma) within 24 hours of blood collection. Upon arrival in the laboratory, blood was continuously rotated at 4 °C before processing to ensure it remained thoroughly mixed. Matched sets of plasma, DBS and PCD were prepared in triplicate from the eighty blood specimens and the complete panels were measured in three different laboratories for α-acid glycoprotein (AGP), C-reactive protein (CRP), Ferritin, histidine rich protein 2 (HRP2), retinol binding protein 4 (RBP4), soluble transferrin receptor (sTfR), and thyroglobulin (Tg) using the MN 7-plex. Because the blood panel was derived from US donors, it was not expected to include malarial parasitemia or a high prevalence of inflammation or iron deficiency [22]. Thus, twenty whole blood samples were randomly chosen from the set and a portion of the whole blood was spiked with calibrator solutions provided by Quansys to simulate samples positive for HRP2 and with elevated AGP, CRP, or sTfR levels. Each contrived sample was prepared by transferring 2 mL of whole blood to a new tube, adding spike solutions and gently mixing by rotating tubes at 4 °C for at least 5 minutes (see Table 1). The resulting samples (Sample numbers 81 – 100) were then processed for DBS and PCD as described below (**Figure 1**).

**Table 1.**
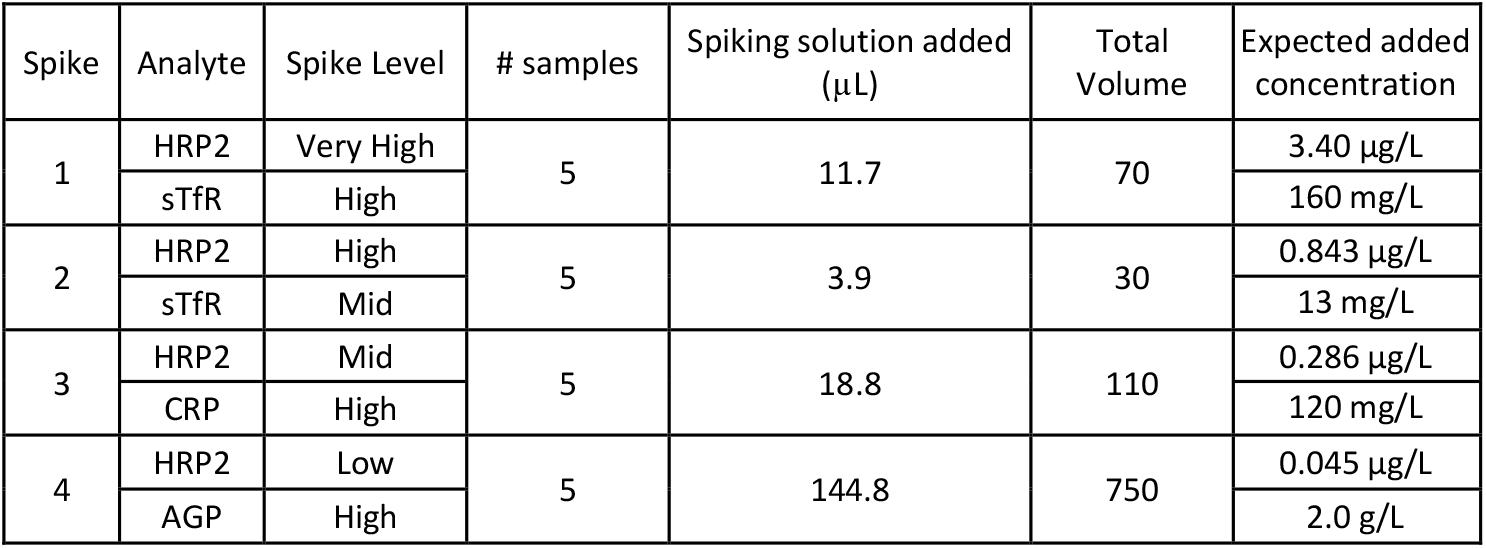
Contrived blood samples spiked with a range of elevated levels of AGP, CRP, HRP2 and sTfR.

**Figure 1.**
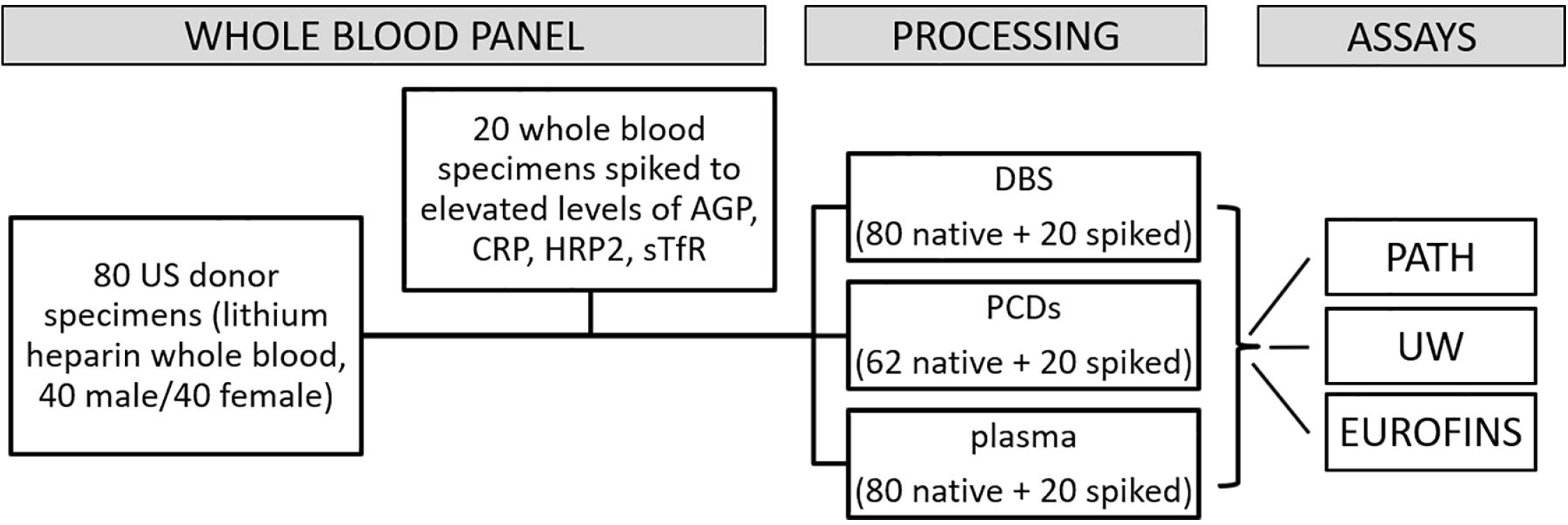
Sample panels and experiment design

DBS were prepared from the 80 whole blood specimens and 20 spiked whole blood specimens by dropping 70 µL of whole blood onto Whatman 903 cards (n=100). The cards were stored overnight to dry at room temperature and then individually stored at -80°C in sealed plastic pouches with desiccant packets.

Prototype PCDs were assembled from parts supplied by the manufacturer for use in these experiments. To produce the PCD, each absorbent plasma collection disc was loaded onto a custom-made manifold and covered by a separation membrane (Figure 2). A volume of 35 µL of whole blood was added to each PCD membrane using a wide-bore pipet tip. After approximately 90 minutes, the PCD membranes containing the RBCs and other cellular components were removed and discarded. The PCDs containing the adsorbed plasma were then removed from the manifold and stored in zipped plastic bags with desiccant at -80 °C. Samples 63-80 were omitted from this portion of the testing for lack of PCD units; the resulting final panel included PCDs from 62 unadulterated donor specimens and 20 PCDs from spiked specimens (n=82). Significant hemolysis was noted on one PCD (ID 12), and corresponding DBS spots appeared very pale. Some slight hemolysis above background was observed on PCDs for 14 samples. Remaining whole blood was then processed to collect plasma fractions (n = 100) as previously described [24]. These plasma fractions were aliquoted into cryovials and stored at -80°C until use. The PCDs were eluted at PATH. The eluted material from the triplicate samples was pooled, mixed and then aliquots of approximately 180 µL was sent to each laboratory on dry ice (see next section), along with matched DBS cards and plasma samples.

**Figure 2.**
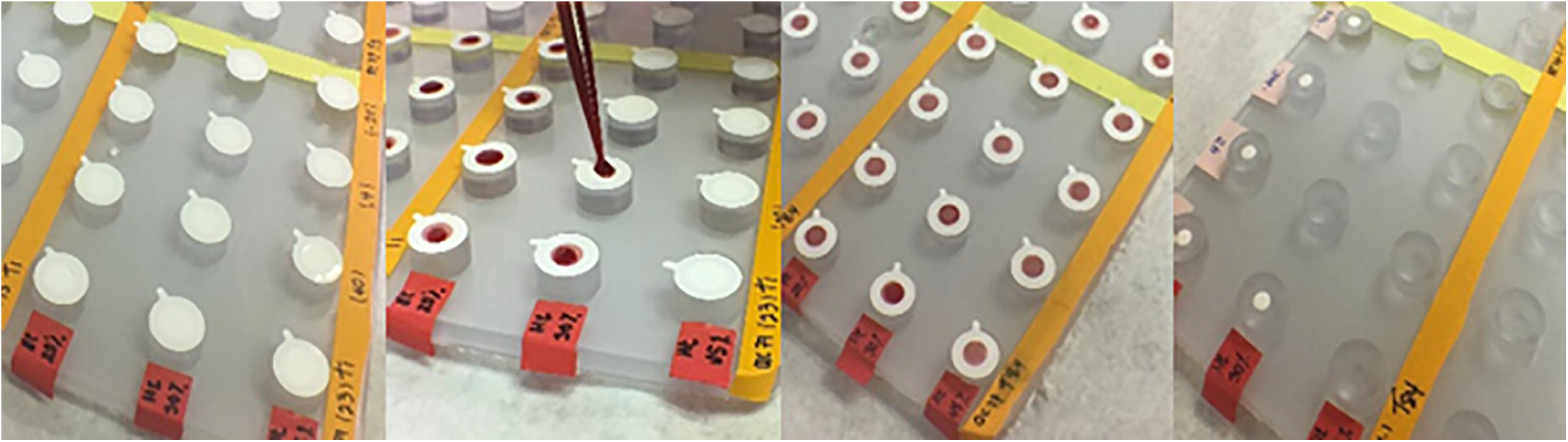
The plasma collection disc in operation. Left to right, the PCD with the the filter disc placed on the manifold prior to whole blood being added, addition of 35 µL of whole blood to each disc, the discs afterblood has been absorebed and finally the plasma collection disc exposed after the whole blood treated filter has been removed. The disc is removed with sterile forceps and is ready for use or can be stored until required.

### Preparation of eluates from DBS and PCDs

The DBS and PCD samples were eluted in the Q-plex assay sample diluent before undergoing analysis with the Q-plex assay. DBS processing was carried out in each laboratory the day before assay. Two 6 mm punches, equivalent to approximately 6 µL of serum [31], were placed in 244 µL sample diluent (approx. 1:20 dilution) and left overnight at 4°C. Samples were then shaken at 200 rpm on a microplate shaker for one hour at room temperature before using. PCD processing occurred in one laboratory, with aliquots of elutes then shared with all 3 laboratories. Each PCD with an estimated equivalent of approximately 8 to 10 µL plasma per PCD was placed in a microcentrifuge tube separation column with a paper filter membrane (Pierce Spin Cups, ThermoFisher Scientific, Waltham, MA, USA) and 200 µL of diluent was added to each PCD, resulting in a dilution of approximately 1 in 20. Samples were incubated at room temperature for two hours before being centrifuged at 12,500 rpm for 15 minutes. Three PCDs per sample were eluted following this procedure, and recovered eluates were pooled for each sample, mixed and aliquoted in ∼180 µL volumes for transfer to the laboratories for testing.

### Human Micronutrient (7-Plex) assay procedure

The MN 7-plex protocol instructs the user to prepare all samples, negative control and calibrator dilutions in sample diluent containing reconstituted competitor. Because the competitor may degrade during the incubation time required for DBS or PCD elution, the standard protocol was modified to add competitor to the mix after sample elution. These protocol alterations were also used for wet plasma specimens included in these analyses.

Competitor was reconstituted in one tenth of the volume of diluent suggested in the MN 7-plex manual to produce a 10X competitor stock and 144 µL of eluate from either DBS or PCD was mixed with 16 µL of 10X assay competitor mix resulting in 1X competitor in a final sample dilution of approximately 1 in 22. Similarly, kit diluent without competitor was used to dilute wet plasma specimens to 1 in 20, and to reconstitute the calibrator provided with the kit and prepare the standard curve; 10X competitor mix was added to prepared solutions as 10% of the final volume. Then 50 µL of each solution was added into the test wells of the Q-plex assay in duplicate, and the remainder of the protocol was carried out as directed in the kit manual.

After addition of the standards and samples, each plate was incubated at room temperature for two hours with shaking at 500 RPM using a flatbed shaker (Titertek Berthold, Huntsville, Alabama, USA). All reactions were aspirated and washed 3 times with 400 µL of wash buffer supplied with the MN 7-plex kit using an automatic plate washer. Next, 50 µL of detection mix was added to each well and the plate was then incubated with shaking at 500 RPM for 1 hour and washed one more time as described above. 50 µL streptavidin horseradish peroxidase solution was added to each well and plates were incubated with shaking for 20 minutes. Following another wash as described above, the chemiluminescent substrate parts A and B were mixed in equal volumes and 50 µL of the mixture was then added to each well. Each plate was then imaged at 270 seconds of exposure time using a Quansys Q-View™ Imager LS (Quansys Biosciences). Q-View™ Software (Quansys Biosciences) was used to overlay a plate map onto the locations of analyte spots in each well and to measure the chemiluminescent signal from each spot in units of pixel intensity. The software applies the calibrator concentration values to the pixel intensities for each spot in the standard curve wells and fits 5 parameter logistic calibration curves for each analyte. The pixel intensities of the spots in each test well are then used to interpolate the concentration of each analyte relative to its calibrator curve. Once the plate image is overlaid with the analysis grid, all of the curve fitting and data reduction steps are automatically applied via the software. The upper and lower limits of quantification determined by Quansys for each kit lot were applied to exclude values beyond the assay concentration ranges. All values were adjusted for dilutions.

### Statistical methods

Reproducibility across the three laboratories was determined using coefficient of variation (CV, mean divided by standard deviation) of the three measures for each sample type. The CVs were also used to assess the inter-assay and inter-laboratory variation. Concordance between AGP, CRP, ferritin, HRP2, RBP4, sTfR, and Tg results in plasma with both DBS (100 pairs) and PCD (82pairs) was evaluated using scatter plots, Lin’s concordance correlation coefficient and Spearman’s correlation coefficient [32]. Agreement in absolute value was evaluated by calculating recovery of plasma values from DBS or PCD (DBS or PCD concentration/plasma concentration, expressed as a percentage) and using Bland Altman plots of the average concentration plotted against the percentage difference in concentrations of each analyte in plasma/DBS or PCD sample pair. Statistical analyses were conducted using Stata 15.1 (StataCorp, College Station, TX, USA). All of the data generated in this study can be publicly accessed at Dataverse (https://doi.org/10.7910/DVN/VY3MDB).

## Results

Summary statistics for each analyte and sample type and CVs for the measures across the three participating laboratories are shown in Table 1. The CVs for plasma and DBS were similar, with higher CVs in the PCDs for most analytes. CVs were extremely high for sTfR, reflecting a large discrepancy between values from one laboratory as compared to those from the other two. Excluding those results reduced CVs, but they remained over 30% for sTfR. CVs were also high for HRP2, regardless of sample type, because of the semi-quantitative nature of the assay. Most specimens in the panel contained no detectable HRP2; spiked samples contained high values near or above the upper limit of detection for the assay. For ferritin, the DBS samples saturated the assay and were at the upper limit of detection, leading to high CVs. This was expected given the presence of red blood cell (RBC) associated ferritin present in this sample type.

Spearman correlations between plasma and DBS and between plasma and PCDs are shown in Table 3. As with Table 2results are shown including and excluding the highly discrepant sTfR data. The Spearman correlation coefficients (Spearman’s rho) between the plasma and the DBS samples were generally high with all being 0.93 or higher with the exception of ferritin. These results closely match previous observations using DBS samples with the 7-plex assay [22]. As expected, the correlation between plasma ferritin and DBS ferritin was low (0.640) as DBS eluates contain a mixture of ferritin from serum and from red blood cells. The correlation of plasma ferritin to PCD ferritin was 0.925, highlighting the potential advantage of PCDs for ferritin measurement in dry samples. The correlation for RBP4 from PCD was lower than from DBS (0.993 versus 0.840). The sTfR correlation for all data was poor (0.545) but was significantly improved when the aberrant data from one site are excluded (0.858).

**Table 2.** Summary statistics and cross-laboratory coefficients of variation (CV) for plasma, DBS and PCD. Spiked samples contain added AGP, CRP, HRP2, and/or sTfR as shown in Table 1. The concentrations are mean values from testing at PATH, UW, and Eurofins. sTfR results are given with and without data from the Eurofins lab. Average CV is the average of the individual sample CVs calculated from results across the 3 laboratories. Abbreviations: AGP, α-1-acid glycoprotein; CRP, C-reactive protein; CV, coefficient of variation; DBS, dried blood spots; N, number; PCD, plasma collection disc; RBP4, retinol-binding protein 4; SD, standard deviation; sTfR, soluble transferrin receptor; Tg, thyroglobulin.

|  |  | Plasma |  |  | DBS |  |  | PCD |  |  |
| --- | --- | --- | --- | --- | --- | --- | --- | --- | --- | --- |
|  |  | N | Mean (SD) | Average CV | N | Mean (SD) | Average CV | N | Mean (SD) | Average CV |
| AGP (g/L) | All | 100 | 1.1 (0.6) | 10.9% | 100 | 1.2 (0.6) | 10.8% | 82 | 0.6 (0.2) | 8.7% |
|  | Unspiked | 80 | 1.0 (0.2) | 11.5% | 80 | 1.1 (0.3) | 11.3% | 62 | 0.5 (0.2) | 7.0% |
|  | Spiked | 20 | 1.55 (1.2) | 8.7% | 20 | 1.7 (1.1) | 9.0% | 20 | 0.7 (0.4) | 14.1% |
| CRP (mg/L) | All | 92 | 7.44 (12.3) | 19.1% | 89 | 5.5 (8.6) | 16.0% | 73 | 2.8 (4.1) | 18.3% |
|  | Unspiked | 73 | 4.93 (5.4) | 20.6% | 70 | 3.6 (4.1) | 18.5% | 54 | 1.9 (2.0) | 20.5% |
|  | Spiked | 19 | 17.08 (22.9) | 13.4% | 19 | 12.2 (15.3) | 7.2% | 19 | 5.3 (7.0) | 13.2% |
| Ferritin (μg/L) | All | 99 | 42.9 (40.6) | 17.7% | 100 | 178.8 (114.1) | 11.6% | 75 | 18.4 (16.0) | 23.6% |
|  | Unspiked | 79 | 43.6 (43.5) | 15.6% | 80 | 178.9 (115.2) | 12.0% | 56 | 19.0 (17.3) | 26.5% |
|  | Spiked | 20 | 39.9 (27.2) | 25.8% | 20 | 178.3 (112.4) | 10.2% | 19 | 16.7 (11.3) | 16.1% |
| HRP2 (μg/L)<br>(semi-quantitative) | All | 26 | 3.59 (4.8) | 18.6% | 100 | 0.79 (2.1) | 60.7% | 49 | 0.9 (1.5) | 52.2% |
|  | Unspiked | 6 | 0.17 (0.1) | 12.2% | 80 | 0.10 (0.0) | 62.2% | 29 | 0.2 (0.0) | 92.6% |
|  | Spiked | 20 | 4.61 (5.1) | 19.0% | 20 | 3.55 (3.5) | 59.4% | 20 | 1.9 (2.0) | 45.8% |
| RBP4 (μmol/L) | All | 100 | 1.67 (0.6) | 12.2% | 100 | 1.20 (0.3) | 9.2% | 82 | 0.6 (0.2) | 10.3% |
|  | Unspiked | 80 | 1.65 (0.6) | 12.7% | 80 | 1.16 (0.3) | 10.4% | 62 | 0.6 (0.2) | 9.5% |
|  | Spiked | 20 | 1.74 (0.6) | 10.5% | 20 | 1.34 (0.4) | 4.4% | 20 | 0.7 (0.2) | 12.6% |
| sTfR (mg/L) | All | 100 | 43.9 (38.6) | 35.3% | 100 | 80.9 (40.2) | 9.2% | 82 | 74.2 (38.2) | 91.9% |
|  | Unspiked | 80 | 40.8 (38.2) | 33.5% | 80 | 79.3 (41.6) | 9.5% | 62 | 69.0 (38.0) | 89.5% |
|  | Spiked | 20 | 56.1 (38.8) | 42.4% | 20 | 87.2 (34.2) | 8.1% | 20 | 90.2 (35.1) | 99.2% |
| sTfR (mg/L),<br>excluding Eurofins | All | 100 | 48.6 (41.3) | 35.9% | 100 | 82.5 (42.0) | 9.9% | 81 | 35.2 (19.8) | 34.3% |
|  | Unspiked | 80 | 44.7 (39.7) | 34.1% | 80 | 80.8 (43.8) | 10.5% | 61 | 33.6 (20.1) | 34.6% |
|  | Spiked | 20 | 64.0 (45.1) | 43.0% | 20 | 89.0 (34.3) | 7.4% | 20 | 39.9 (18.7) | 33.2% |
| Tg (μg/L) | All | 98 | 23.8 (22.1) | 12.2% | 99 | 21.1 (21.7) | 8.7% | 81 | 12.4 (13.3) | 27.5% |
|  | Unspiked | 78 | 22.6 (20.5) | 13.1% | 79 | 20.0 (19.6) | 9.2% | 61 | 11.2 (10.7) | 28.0% |
|  | Spiked | 20 | 28.4 (27.7) | 8.9% | 20 | 25.5 (28.9) | 6.7% | 20 | 16.1 (19.2) | 26.2% |

**Table 3.** The comparison of results for plasma, DBS and PCD eluates using Spearman correlation and recovery of expected plasma values. For the PCD, the adjusted values reflect an estimate of 5 µL of plasma from each disc instead of 10 µL of plasma as originally estimated. For sTfR, results are shown with and without data from Eurofins. Abbreviations: AGP, α-1-acid glycoprotein; CRP, C-reactive protein; DBS, dried blood spots; EF, Eurofins ; N, number; PCD plasma collection disc; RBP4, retinol-binding protein 4; SD, standard deviation; sTfR, soluble transferrin receptor; Tg, thyroglobulin. Rho, rank-order correlation.

|  | AGP | CRP | Ferritin | HRP2 | RBP4 | sTfR | sTfR<br>excl. EF | Tg |
| --- | --- | --- | --- | --- | --- | --- | --- | --- |
| DBS |  |  |  |  |  |  |  |  |
| N (pairs) | 100 | 89 | 99 | 26 | 100 | 100 | 100 | 98 |
| Spearman's Rho | 0.975 | 0.988 | 0.640 | 0.977 | 0.993 | 0.930 | 0.947 | 0.995 |
| Recovery, mean (SD%) | 116 (15) | 71 (14) | 570<br>(400) | 98 (55) | 74 (9) | 234 (80) | 215 (76) | 89 (11) |
| PCD |  |  |  |  |  |  |  |  |
| N (pairs) | 82 | 71 | 75 | 21 | 82 | 82 | 81 | 81 |
| Spearman's Rho | 0.954 | 0.981 | 0.925 | 0.919 | 0.840 | 0.583 | 0.858 | 0.977 |
| Recovery mean (SD%) | 51 (11) | 39 (18) | 45 (15) | 52 (33) | 37 (8) | 218<br>(110) | 88 (35) | 51 (12) |
| Adj. Spearman's Rho | 0.835 | 0.965 | 0.848 | 0.914 | 0.760 | 0.798 | 0.855 | 0.964 |
| Adj. Recovery mean<br>(SD) % | 101 (21) | 79 (36) | 90 (29) | 103 (66) | 73 (16) | 436<br>(220) | 176 (69) | 102 (24) |

The mean recovery compares concentrations of each analyte in DBS or PCDs relative to plasma concentrations (Table 3). Concentrations of all, spiked, and unspiked, samples were similar for plasma and DBS, but PCD values were approximately half the values measured in plasma. For AGP, HRP2 and Tg, values recovered from DBS were within one standard deviation of 100% of the plasma values, while CRP and RBP4 recoveries from DBS were lower at 71% and 74%, respectively. For DBS versus plasma sTfR, the recovery rates from DBS were very high at 234%. Excluding results from one of the three sites only marginally improved DBS recovery relative to plasma (215%) The impact of the discrepant sTfR data were more pronounced for the recovery from PCDs relative to plasma; including all data, the sTfR recovered from PCDs was 436% ±220% relative to plasma, but when sTfR results from the Eurofins lab are excluded, the PCD values were 176% ± 69% of plasma.

Original concentration estimates for the PCD eluates used the assumption that each disc contained the equivalent of approximately 10 µL of plasma, resulting in a dilution of 1 in 22 when the disc was eluted. Recovery results shown in Table 3 suggest that the volume of plasma recovered from the disc is approximately 50% of the original estimate, giving an eluate dilution of 1 in 44. Adjusting the PCD results to account for the new plasma volume equivalent improved agreement with plasma values in terms of absolute values, but had minimal impacts on correlation (Table 3, Figure 3Aand 3B).

**Figure 3A.**
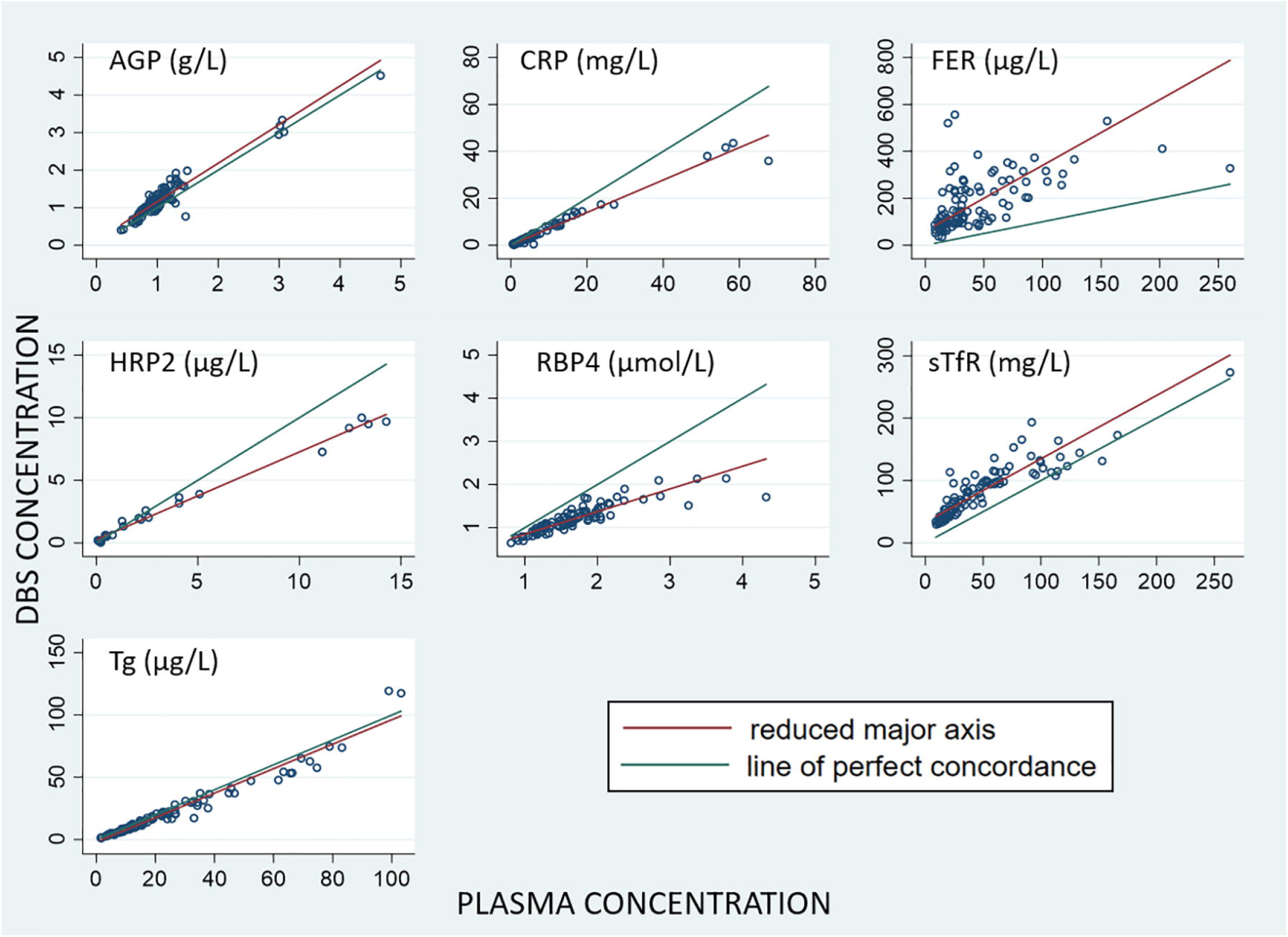
Lin’s CCC plots comparing the analyte measurements in paired wet plasma (x-axes) and DBS samples (y-axes).

**Figure 3B.**
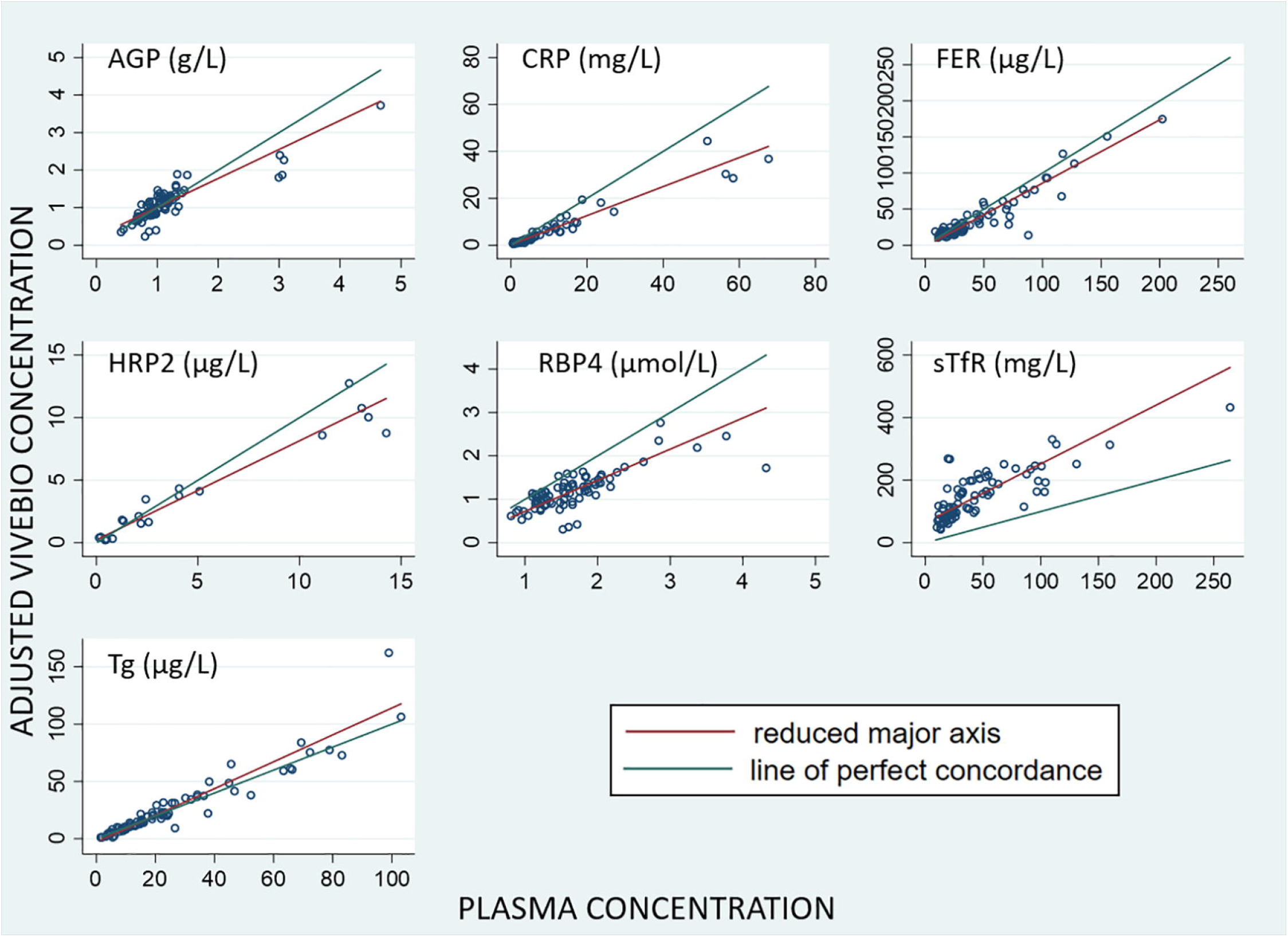
Lin’s CCC plots comparing the analyte measurements in paired wet plasma (x-axes) and the adjusted PCD (, y-axes). AGP, α-1-acid glycoprotein; CRP, C-reactive protein; FER, ferritin; HRP2, histidine rich protein 2; RBP4, retinol binding protein 4; sTfR, soluble transferrin receptor; Tg, thyroglobulin; DBS, dried blood spot.

To complete our comparative analysis we also assessed the correlation of the DBS and the adjusted PCD samples to the plasma samples using scatter plots with linear regression lines (Figures 3A and 3B) and their agreement using Bland Altman plots (Figures 4A and 4B). We used Lin’s concordance correlation coefficient (CCC) to create scatter plots to compare the measurements [33]. AGP and Tg were both very close to the line of perfect concordance and sTfR is correlative though the reduced major axis is consistently greater than the line of perfect correlation. HRP2, CRP and RBP4 are less concordant while bias increases showing greater variance at the higher analyte concentrations. The plot for ferritin shows the lowest correlation as is expected since DBS samples have ferritin contributed from RBCs. With the exception of the sTfR plot, all other DBS analytes tended to be lower than plasma values. For most analytes, the correlation of adjusted PCD and plasma was similar to that of DBS and plasma.

**Figure 4A.**
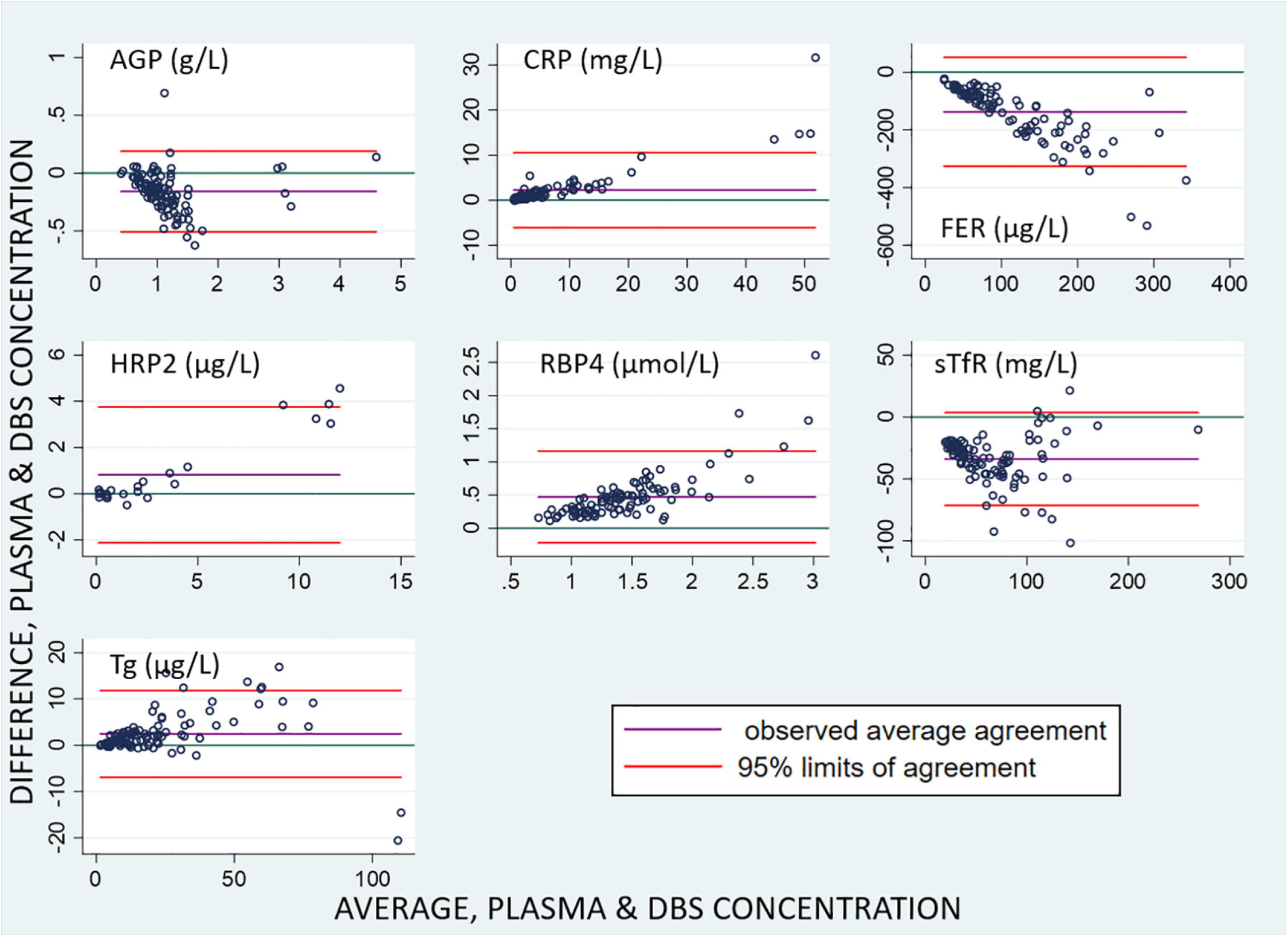
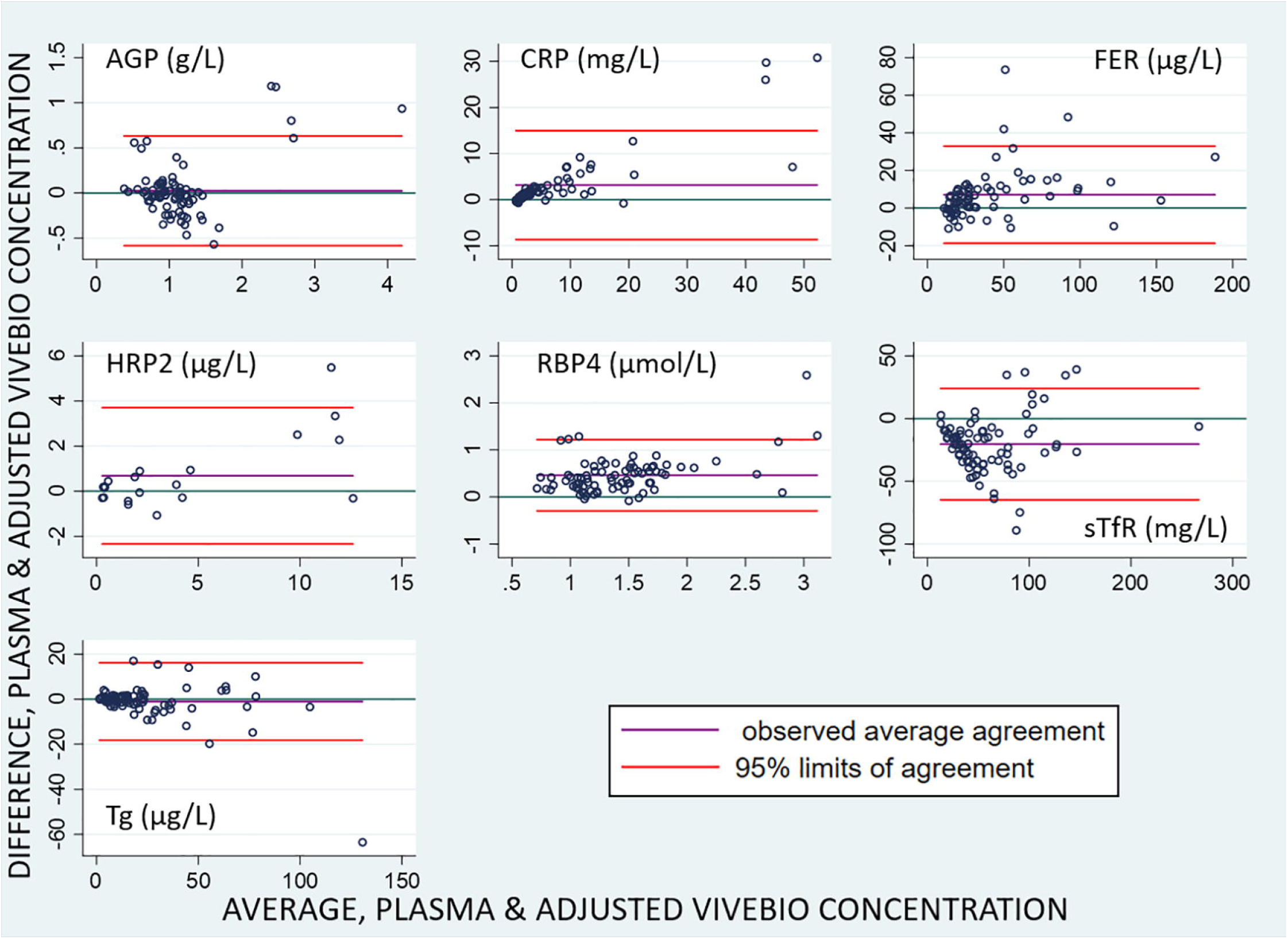
Plots of the difference in results derived from wet plasma versus dry samples types measured using the 7-plex assay. Average of the wet plasma and dry sample type (x-axes) are plotted against the difference between measurements from the 7-Plex assay for paired wet plasma and dry sample types (y-axes). Fig 3A, the plasma versus DBS samples; **Figure 4B**. the plasma versus the adjusted PCD. Horizontal lines indicate line of perfect agreement (green), mean (purple), and ± 2standard deviations of the difference (red). AGP, α-1-acid glycoprotein; CRP, C-reactive protein; FER, ferritin; HRP2, histidine rich protein 2; RBP4, retinol binding protein 4; sTfR, soluble transferrin receptor; Tg, thyroglobulin; DBS, dried blood spot.

## Discussion

Correlation, concordance plots, and Bland Altman plots suggest that plasma and PCDs show good agreement for AGP, CRP, ferritin, HRP2, RBP, and Tg, but not sTfR. The sTfR results show gross variance between the absolute values for all three sample types (see Table 3). The sTfR assay remained the least concordant, especially at the higher values. The increasing bias was again evident with the samples having more variance at the higher concentrations of analyte. Most of the plots (Fgirues 3 and 4) showed a slight positive bias at the upper dynamic ranges of each assay. With the exception of sTfR and marginally HRP2, the trend was that the values for the adjusted PCD were below those for plasma. As expected, the agreement of PCD data for the ferritin data sets was signifcantly better than with the DBS samples. The primary advantage of PCDs over DBS is the improved correlation of ferritin results from plasma and dry blood with the removal of RBCs from the samples.

One significant limitation that was identified was uncertainty about the volume of plasma captured in each disc; this needs to be fully validated. We had estimated that each PSC absorbed a volume of 10 µL plasma per disc but it was evident from the mean recoveries that the discs either contained ∼50% less plasma or that some material was bound or retained within the disc. Therefore a rigorous assessment of the volumetric capacity of the PCD is warranted, essentially mimicking the effort to assess the volume of material retained in DBS relative to sampling volumes [20]. Earlier work on dried serum spots spotted precise volumes of serum onto filter paper and so the exact volume of material was known, simplifying the harmonization of data. We suspect that while processing PCD with whole blood under vacuum, some of the serum/plasma was drawn from the disc and lost, resulting in smaller sample volume being retained in the dead space of the disc pores. It is also possible that DriVive production version of the PCDs will retain plasma more efficiently than did our prototype versions assembled in the lab.

In this work, the samples were eluted from the PCDs soon after preparation and stored at low temperatures to preserve the integrity of the samples before testing. Future tests will be required to understand the stability of the biomarkers in dried PCDs as their intended use will include population based micronutrient surveys in austere settings where access to refrigeration will be limited or absent, so adequate stability of the samples out of cold chain is essential. Dried serum spots have been shown to be stable for at least two weeks but in this case the precise volume of serum per serum spot was known and so comparative analyses is easier [34].

The sTfR data is a concern in that the original DBS and PCD samples had mean recoveries of over 200%, due in part to elevated sTfR from one partner laboratory. The source of this discrepancy could not be identified; all laboratories used the same lot of plates and the other 6 assays in the multiplex gave mean recoveries similar to the paired data from other laboratories meaning the issue was specific to the sTfR assay performed in the one laboratory. When this data was removed to create the adjusted datasets, the mean recovery was not significantly changed with DBS samples. With the exclusive PSC data the sTfR recovery did drop significantly but when the adjustment for volumetric bias was applied the mean recovery was still high at 176 mg/mL. Since we conducted this evaluation, Quansys have released a new version of MN 7-plex array in which they claim the sTfR assay has been completely redeveloped with new antibodies. Quansys reports that the new assay has significantly better performance when assessed with the Vital EQA panel provided by the US CDC [35]. We are currently assessing the performance of the previous and new version of the array to independently confirm that the discordance and elevated recoveries observed with sTfR are significantly reduced.

From this study it appears that the PCD has potential to be an effective tool for the rapid instrument free fractionation of plasma from whole blood samples. Previously, we had shown that DBS could be used as a specimen type for use with the MN-7 plex assay, but DBS are limited to only 6 of the biomarkers (AGP, CRP, HRP2, RBP4, sTfR and Tg) [22]. The inability to accurately measure serum ferritin is a significant drawback to using DBS if measuring iron levels is a primary goal, as sTfR measurement alone is insufficient for accurate determination of iron deficiency. Here we have shown that ferritin can be measured from the PCD and that the values derived were correlative to and in agreement with plasma samples and with other previously reported work [34,36]. In this experiment, we were able to evaluate HRP2 as an indicator of *Plasmodium falciparum* antigenemia via spiking with the antigen, thus adding an assessment missing from our previous test of the use of DBS with the MN 7-plex. HRP2 was recovered well from both DBS and the PCD eluates, suggesting that the malarial antigen assay is functional with either DBS or PCD samples. Thus, with the addition of this evidence about the performance of HRP2 and ferritin measures in samples collected using PCDs, our results suggest that all seven analytes in the MN 7-plex can be measured reliably using a dry sample collection method that eliminates the need for sample processing in the field and minimizes cold chain requirements.

## Data Availability

All data produced are available online at once the manuscript has been approved for publication.

https://doi.org/10.7910/DVN/VY3MDB

## Acknowledgments

We greatly appreciate the collaborative support of ViveBio LLC in supplying prototype PCD devices and their expert technical input. We would like to thank Dr Emily Smith (The George Washington University) and Dr Ken Brown (U.C. Davis) for their critical thinking and advice. We would also like to thank Ms. Olivia Halas for her administrative support in preparing and reviewing the manuscript for submission.

